# Trends in, and factors associated with, chemsex in a community sample of gay, bisexual, and other men who have sex with men in England, 2022-2025

**DOI:** 10.64898/2026.10.19.26365930

**Authors:** Jin-Min Yuan, Laura Viviani, Dolores Mullen, Dana Ogaz, Hamish Mohammed

## Abstract

**Background:** Chemsex, drug use before/during sex to facilitate and enhance the experience, is undertaken by a minority of gay, bisexual and other men who have sex with men (GBMSM), but evidence of chemsex prevalence in England is limited.

This study describes trends in chemsex prevalence between 2022-2025 and identifies factors associated with chemsex amongst GBMSM in England.

**Methods:** Data from the RiiSH study (2022-2025), an annual online cross-sectional survey of GBMSM recruited through social networking/dating apps and community partners, were analysed.

Trends in chemsex prevalence and adjusted prevalence were estimated using log-binomial regression models.

The following seven variables were candidate factors associated with chemsex and were assessed through logistic regression: age, sexual orientation, education, employment, HIV status, HIV PrEP use, and sexual health service (SHS) use.

**Results:** 5728 GBMSM were included in this study. Adjusted chemsex prevalence varied from 6.1% in 2022 to 4.2% in 2023, 5.1% in 2024 and 6.8% in 2025.

Chemsex was more likely in GBMSM who were: gay (aOR=1.57 versus other sexual orientations; 95%CI 1.11-2.29, p=0.01), unemployed (aOR=3.08 versus employed full-time; 95%CI 2.19-4.26, p<0.001); aged 35-59 versus 16-34 years (p<0.001), with recent HIV PrEP use (aOR=3.79 versus no recent use; 95%CI 2.63-5.56, p<0.001), SHS use (aOR=2.23 versus no recent use; 95%CI 1.76-2.83, p<0.001), and living with HIV (aOR=3.16 versus negative/unknown HIV status; 95%CI 2.43-4.09, p<0.001).

**Conclusions:** Chemsex prevalence was low and stable between 2022-2025. PrEP and SHS use were associated with chemsex, supporting integration of specialist support services within SHS.

## Introduction

Chemsex, the use of crystal methamphetamine (‘crystal meth’), mephedrone and gamma-hydroxybutyrate/gamma-butyrolactone (GHB/GBL) before or during planned sexual events to facilitate, enhance, prolong and/or sustain the experience, is a practice undertaken by a minority of gay, bisexual and other men who have sex with men (GBMSM) (McCall et al., 2015; Melendez-Torres et al., 2018).

Although labelling drug use as problematic can be unhelpful (Stuart, 2019), and notwithstanding the structures of care and harm reduction that already exist within GBMSM networks (Pienaar et al., 2018), it is recognised that chemsex can lead to significant health harms. These include: sexually transmitted infection (STI) transmission (Flores Anato et al., 2022; Kohli et al., 2019; MacGregor et al., 2021), skin and soft tissue injuries from injecting chemsex drugs (‘slamming’) (Bourne et al., 2015), and overdoses – particularly of GHB/GBL – leading to loss of consciousness, non-consensual sex (Drückler et al., 2021), and death in some instances (Bourne et al., 2015). Positive impacts of chemsex have been reported, with higher measures of sexual self-esteem and satisfaction associated with its use (Demant et al., 2025). Nevertheless, chemsex can also impact negatively on mental and social wellbeing (Berg et al., 2020; Tomkins et al., 2019), with damage to social relationships and work careers as a result of prioritising chemsex (Bourne et al., 2015).

Previous research has demonstrated associations between chemsex/chemsex-associated drug use and sociodemographic characteristics (Adler et al., 2022; Blomquist et al., 2020; Demant et al., 2025; Frankis et al., 2018; Gonzalez-Recio et al., 2024; Guerras et al., 2021; Hammond et al., 2023; Hibbert et al., 2019; Jalil et al., 2022; Rodríguez-Expósito et al., 2024; Schmidt et al., 2016; Sewell et al., 2017), relationship status (Demant et al., 2025; Frankis et al., 2018; Malandain et al., 2021; Sewell et al., 2017), education and employment (Connolly et al., 2023; Hammond et al., 2023; Ly et al., 2025; Paudel et al., 2025; Sewell et al., 2017), geographical residence (Gonzalez-Recio et al., 2024; Guerras et al., 2021, 2022; Hibbert et al., 2019; Jalil et al., 2022; Schmidt et al., 2016; Sewell et al., 2017), drug and alcohol use (Elin et al., 2025; Evers et al., 2019; Gonzalez-Recio et al., 2024; Hammond et al., 2023; Hibbert et al., 2019; Ivey et al., 2023; Jalil et al., 2022; Ly et al., 2025; Maviglia et al., 2022; Rosas Cancio-Suárez et al., 2023; Sewell et al., 2017), unprotected anal sex (Chone et al., 2021; Eustaquio et al., 2024a; Glynn et al., 2018; González-Baeza et al., 2018; Guerras et al., 2021, 2022; Hammond et al., 2023; Ivey et al., 2023; Jalil et al., 2022; Pufall et al., 2018; Sewell et al., 2017), transactional sex (Frankis et al., 2018; Guerras et al., 2021, 2022; Ly et al., 2025; Maviglia et al., 2022), sexually transmitted infections (Adler et al., 2022; Amundsen et al., 2022; Capodieci et al., 2025; Drückler et al., 2018; Eustaquio et al., 2024b; Glynn et al., 2018; González-Baeza et al., 2018; Guerras et al., 2021, 2022; Hammond et al., 2023; Hampel et al., 2020; Hibbert et al., 2019; Jalil et al., 2022; Lee et al., 2021; Maviglia et al., 2022; Pufall et al., 2018; Sewell et al., 2017; Wong et al., 2025a), use of HIV pre-exposure prophylaxis (PrEP) use (Chone et al., 2021; Jalil et al., 2022; Wong et al., 2025a) and mental ill health (Connolly et al., 2023; Demant et al., 2025; Hampel et al., 2020; Ivey et al., 2023; Paudel et al., 2025; Sewell et al., 2017; Wong et al., 2025b). However, these studies have been undertaken in different time periods and multiple different countries, where the societal context and healthcare service may differ significantly from England.

We used data from four rounds of an annual online cross-sectional survey of GBMSM to estimate the prevalence of chemsex in England in 2022-2025. We also aimed to identify factors associated with recent chemsex specific to the population and healthcare context in England.

## Methods

### Study design and data sources

The ‘Reducing inequalities in Sexual Health’ (RiiSH) survey is a self-completed annual online cross-sectional survey generally run between November-December recruiting self-identifying men (cisgender/transgender), transgender women, or gender-diverse people assigned male at birth, who are aged 16 years or over, resident in the UK, and report having had sex with a man (cisgender and/or transgender) in the last year. This is a convenience sample with participants recruited through social networking sites (Facebook, Instagram), geospatial dating platforms (Grindr, Scruff, Jack’d, Recon) and community organisations.

Analyses in this study were restricted to participants identifying as men, due to the small numbers of participants identifying as transgender women or gender-diverse, and those resident in England because sexual health commissioning differs across the UK.

The RiiSH survey asks questions related to sociodemographic characteristics, sexual health behaviours, health service interactions and drug use. Some questions refer to the period covering the 3-4 months prior to survey completion; we labelled any behaviours reported in this timeframe as ‘recent’.

### Outcome of interest

Recent chemsex was the outcome of interest, defined as any sex after taking crystal meth, mephedrone or GHB/GBL in the last 3-4 months.

### Statistical analysis

RiiSH survey responses from 2022-2025 were extracted and cleaned.

#### 1) Trends in chemsex prevalence

Trends in chemsex prevalence over time were assessed using multivariable log-binomial regression to estimate adjusted risk ratios (aRR). The proportion of participants recruited to the RiiSH survey by each recruitment source varies by year, and participants recruited via dating (as opposed to social media) apps are more likely to report STI risk behaviours (Bezerra et al., 2022; Cabecinha et al., 2017; Gibson et al., 2022; Lewnard & Berrang-Ford, 2014). Given the potential clustering of participants within recruitment sources, we included recruitment source as a covariate in the final model.

The RiiSH survey in 2022 was publicised widely to assess service use, sexual risk behaviour and mpox vaccination uptake after the mpox outbreak in England that summer. As a result, the participant demographics may have been particularly different in 2022 than in subsequent years in relation to sexual health risk. To assess the impact of this, in a sensitivity analysis we additionally adjusted for the number of condomless anal intercourse partners participants reported having sex with in the 3-4 month period prior to survey completion as a measure of sexual risk.

Adjusted chemsex prevalence estimates for each survey year were obtained from marginal predictions generated from the fitted model.

#### 2) Factors associated with chemsex

To facilitate a theory-driven approach, a rapid literature review of known factors associated with chemsex (see search strategy in Appendix 1) and discussions with subject matter experts at the UK Health Security Agency (UKHSA) were undertaken to inform the *a priori* shortlisting of the following variables, each of which have a potential role in the use of chemsex (see appendix 2 for the full list): age, sexual orientation, education level, employment status, HIV status, recent HIV PrEP use, and recent sexual health service (SHS) use.

A directed acyclic graph (DAG) was built in DAGitty (Textor et al., 2011) to represent associations between chemsex use and the variables, informed by the rapid literature review and subject matter expert discussion (Appendix 3). Logistic regression models were fitted to estimate unadjusted and adjusted odds ratios for chemsex use. The multivariable model for each variable of interest was adjusted for age (as a baseline covariate) and the minimally-sufficient adjustment set identified by the DAG. This approach facilitated the careful selection of confounders, minimised overadjustment and increased the transparency of our regression models (Gao et al., 2025; Lewer et al., 2025).

All analyses were undertaken in RStudio 2026.07.0 (Posit team, 2026).

### Ethical approval

Ethical approval for this study was granted by the UKHSA Research Ethics and Governance Group (reference R&D 524).

## Results

### Descriptive analyses

Overall, 5728 RiiSH participants from the 2022-2025 RiiSH rounds were included in this study (Table 1). Most participants were recruited from geospatial networking apps (67%) or social media (27%). The modal age-group was 45-59 years (37%), followed by 35-44 years (25%). Most participants were of White ethnicity (88%) and identified as gay (83%).

**Table 1:** Characteristics of participants from the Reducing inequalities in Sexual Health survey undertaken by gay, bisexual, and other men who have sex with men, England, 2022-2025, stratified by whether they reported recent chemsex (in the last 3-4 months). *For ethnicity, some strata have been combined in this table to protect participants’ privacy

| Characteristic | Recent chemsex |  |  |
| --- | --- | --- | --- |
|  | Overall<br>N = 5,728<br>n | No<br>N = 5,407<br>n (%) | Yes<br>N = 321<br>n (%) |
| **Survey year** |  |  |  |
| 2022 | 1,111 | 1,038 (93%) | 73 (6.6%) |
| 2023 | 892 | 853 (96%) | 39 (4.4%) |
| 2024 | 2,272 | 2,161 (95%) | 111 (4.9%) |
| 2025 | 1,453 | 1,355 (93%) | 98 (6.7%) |
| **Recruitment source** |  |  |  |
| community/other | 385 | 356 (92%) | 29 (7.5%) |
| geospatial networking app | 3,815 | 3,617 (95%) | 198 (5.2%) |
| social media | 1,528 | 1,434 (94%) | 94 (6.2%) |
| **Age (years)** |  |  |  |
| 16-34 | 1,217 | 1,175 (97%) | 42 (3.5%) |
| 35-44 | 1,424 | 1,323 (93%) | 101 (7.1%) |
| 45-59 | 2,115 | 1,983 (94%) | 132 (6.2%) |
| 60+ | 972 | 926 (95%) | 46 (4.7%) |
| **Ethnicity** |  |  |  |
| White | 5,067 | 4,777 (94%) | 290 (5.7%) |
| Asian, Asian British or Asian Welsh | 273 | 264 (97%) | 9 (3.3%) |
| Black, Black British, Black Welsh, Caribbean or African/Other ethnic group | 215 | 207 (96%) | 8 (3.7%) |
| Mixed or Multiple ethnic groups | 173 | 159 (92%) | 14 (8.1%) |
| **Sexual orientation** |  |  |  |
| Bisexual/Heterosexual/Other/Prefer not to say | 959 | 924 (96%) | 35 (3.6%) |
| Gay or homosexual | 4,769 | 4,483 (94%) | 286 (6.0%) |
|  | Overall<br>N = 5,728<br>n | No<br>N = 5,407<br>n (%) | Yes<br>N = 321<br>n (%) |
| **Financial situation** |  |  |  |
| Financially comfortable | 2,616 | 2,495 (95%) | 121 (4.6%) |
| Financially ok | 1,639 | 1,559 (95%) | 80 (4.9%) |
| Financially uncomfortable | 1,360 | 1,244 (91%) | 116 (8.5%) |
| Unknown | 113 | 109 | 4 |
| **Education level** |  |  |  |
| Below degree-level | 2,017 | 1,918 (95%) | 99 (4.9%) |
| Degree or higher | 3,488 | 3,276 (94%) | 212 (6.1%) |
| No qualifications/other | 223 | 213 (96%) | 10 (4.5%) |
| **Employment status** |  |  |  |
| Employed full time | 4,049 | 3,842 (95%) | 207 (5.1%) |
| Employed part time | 398 | 378 (95%) | 20 (5.0%) |
| Student | 188 | 183 (97%) | 5 (2.7%) |
| Unemployed/Other | 391 | 340 (87%) | 51 (13%) |
| Retired/disabled/long-term sickness | 702 | 664 (95%) | 38 (5.4%) |
| **Recent injecting drug use** |  |  |  |
| No | 5,637 | 5,394 (96%) | 243 (4.3%) |
| Yes | 91 | 13 (14%) | 78 (86%) |
| **Recent use of HIV PrEP** |  |  |  |
| No (including people living with HIV) | 3,266 | 3,127 (96%) | 139 (4.3%) |
| Yes | 2,444 | 2,263 (93%) | 181 (7.4%) |
| Unknown | 18 | 17 | 1 |
| **HIV status** |  |  |  |
| HIV negative/unknown | 4,989 | 4,766 (96%) | 223 (4.5%) |
| Living with HIV | 739 | 641 (87%) | 98 (13%) |
| **Recent sexual health service (SHS) use** |  |  |  |
| No | 3,351 | 3,227 (96%) | 124 (3.7%) |
| Yes | 2,372 | 2,177 (92%) | 195 (8.2%) |
|  | Overall<br>N = 5,728<br>n | No<br>N = 5,407<br>n (%) | Yes<br>N = 321<br>n (%) |
| Unknown | 5 | 3 | 2 |
| **Recent condomless anal intercourse (CAS)** |  |  |  |
| No | 1,836 | 1,808 (98%) | 28 (1.5%) |
| Yes | 3,892 | 3,599 (92%) | 293 (7.5%) |
| **Number of recent (past 3-4 months) CAS partners** |  |  |  |
| 0 | 1,836 | 1,808 (98%) | 28 (1.5%) |
| 1 | 1,092 | 1,082 (99%) | 10 (0.9%) |
| 2-4 | 1,405 | 1,338 (95%) | 67 (4.8%) |
| 5-9 | 626 | 552 (88%) | 74 (12%) |
| 10+ | 769 | 627 (82%) | 142 (18%) |

Overall, recent chemsex was reported by 5.6% (321/5728) of study participants, with some variation by recruitment source: recent chemsex was reported by 7.5% (29/385) of community/other-recruited participants, 6.2% (94/1528) of social media-recruited participants and 5.2% (198/3815) of geospatial networking app-recruited participants.

Recent PrEP and SHS use were both reported by over 40% of participants. Two thirds of participants reported recent condomless anal intercourse, of which 72% had more than one recent condomless anal intercourse partner. A minority reported recent injecting drug use (1.6%), but of these 86% reported recent chemsex.

### Trends in recent chemsex prevalence

Recent chemsex prevalence varied between 2022 to 2025: relative to 2022, it was lower in 2023 (aRR 0.70; 95%CI 0.47-1.03; p=0.07) and 2024 (aRR 0.84; 95%CI 0.60-1.20; p=0.3), and similar in 2025 (aRR 1.13; 95%CI 0.78-1.64; p=0.5), in both univariable and multivariable models (Table 2).

**Table 2:** Univariable and multivariable analysis of chemsex prevalence by Reducing inequalities in Sexual Health survey year, among gay, bisexual, and other men who have sex with men, England, 2022-2025. *Adjusted for age and recruitment source

| Variable | Unadjusted risk ratio | 95% CI | p-value | Adjusted risk ratio* | 95% CI | p-value |
| --- | --- | --- | --- | --- | --- | --- |
| Year |  |  | 0.02 |  |  | 0.04 |
| 2022 | — | — |  | — | — |  |
| 2023 | 0.67 | 0.45, 0.97 | 0.04 | 0.70 | 0.47, 1.03 | 0.07 |
| 2024 | 0.74 | 0.56, 0.99 | 0.04 | 0.84 | 0.60, 1.20 | 0.3 |
| 2025 | 1.03 | 0.77, 1.38 | 0.9 | 1.13 | 0.78, 1.64 | 0.5 |

**Table 3:** Univariable and multivariable analyses of factors associated with recent chemsex, among gay, bisexual, and other men who have sex with men, England, 2022-2025. *Covariates included in the multivariable models for each independent variable are outlined in Appendix 3. For age, no confounders were identified so only the unadjusted model coefficients are presented here.

| Independent variables | N | Unadjusted odds ratio | 95% CI | p-value | Adjusted odds ratio* | 95% CI | p-value |
| --- | --- | --- | --- | --- | --- | --- | --- |
| Age (years) | 5,728 |  |  | <0.001 |  |  |  |
| 16-34 |  | — | — |  |  |  |  |
| 35-44 |  | 2.14 | 1.49, 3.12 | <0.001 |  |  |  |
| 45-59 |  | 1.86 | 1.32, 2.68 | <0.001 |  |  |  |
| 60+ |  | 1.39 | 0.91, 2.14 | 0.13 |  |  |  |
| Sexual orientation | 5,728 |  |  | 0.002 |  |  | 0.01 |
| Bisexual/Heterosexual/Other/Prefer not to say |  | — | — |  | — | — |  |
| Gay or homosexual |  | 1.68 | 1.19, 2.45 | 0.004 | 1.57 | 1.11, 2.29 | 0.01 |
| Education level | 5,728 |  |  | 0.14 |  |  | 0.13 |
| Degree or higher |  | — | — |  | — | — |  |
| Below degree-level |  | 1.25 | 0.98, 1.61 | 0.07 | 1.26 | 0.99, 1.62 | 0.06 |
| No qualifications/other |  | 0.91 | 0.44, 1.69 | 0.8 | 0.93 | 0.45, 1.74 | 0.8 |
| Employment status | 5,728 |  |  | <0.001 |  |  | <0.001 |
| Employed full time |  | — | — |  | — | — |  |
| Employed part time |  | 0.98 | 0.60, 1.53 | 0.94 | 1.13 | 0.68, 1.78 | 0.6 |
| Student |  | 0.51 | 0.18, 1.12 | 0.14 | 0.76 | 0.26, 1.74 | 0.6 |
| Unemployed/Other |  | 2.78 | 1.99, 3.83 | <0.001 | 3.08 | 2.19, 4.26 | <0.001 |
| Retired/disabled/long-term sickness |  | 1.06 | 0.73, 1.50 | 0.7 | 1.33 | 0.86, 2.02 | 0.2 |
| Recent HIV PrEP use | 5,710 |  |  | <0.001 |  |  | <0.001 |
| No PrEP (including people living with HIV) |  | — | — |  | — | — |  |
| Yes PrEP |  | 1.80 | 1.43, 2.26 | <0.001 | 3.79 | 2.63, 5.56 | <0.001 |
| HIV status | 5,728 |  |  | <0.001 |  |  | <0.001 |
| HIV negative/unknown |  | — | — |  | — | — |  |
| Living with HIV |  | 3.27 | 2.53, 4.19 | <0.001 | 3.16 | 2.43, 4.09 | <0.001 |
| Recent sexual health service use | 5,723 |  |  | <0.001 |  |  | <0.001 |
| No |  | — | — |  | — | — |  |
| Yes |  | 2.33 | 1.85, 2.94 | <0.001 | 2.23 | 1.76, 2.83 | <0.001 |

Abbreviations: CI = Confidence Interval, OR = Odds Ratio

Adjusted chemsex prevalence estimates fluctuated, from 6.1% (95%CI 4.5-7.7) in 2022 to 4.2% (95%CI 2.9-5.6) in 2023, 5.1% (95%CI 4.1-6.1) in 2024 and 6.8% (95%CI 5.4-8.3) in 2025 (Figure 1). The same pattern was observed in sensitivity analysis, where we additionally adjusted for the number of condomless anal intercourse partners participants reported having sex with in the 3-4 months prior to survey completion (Appendix 4).

**Figure 1:**
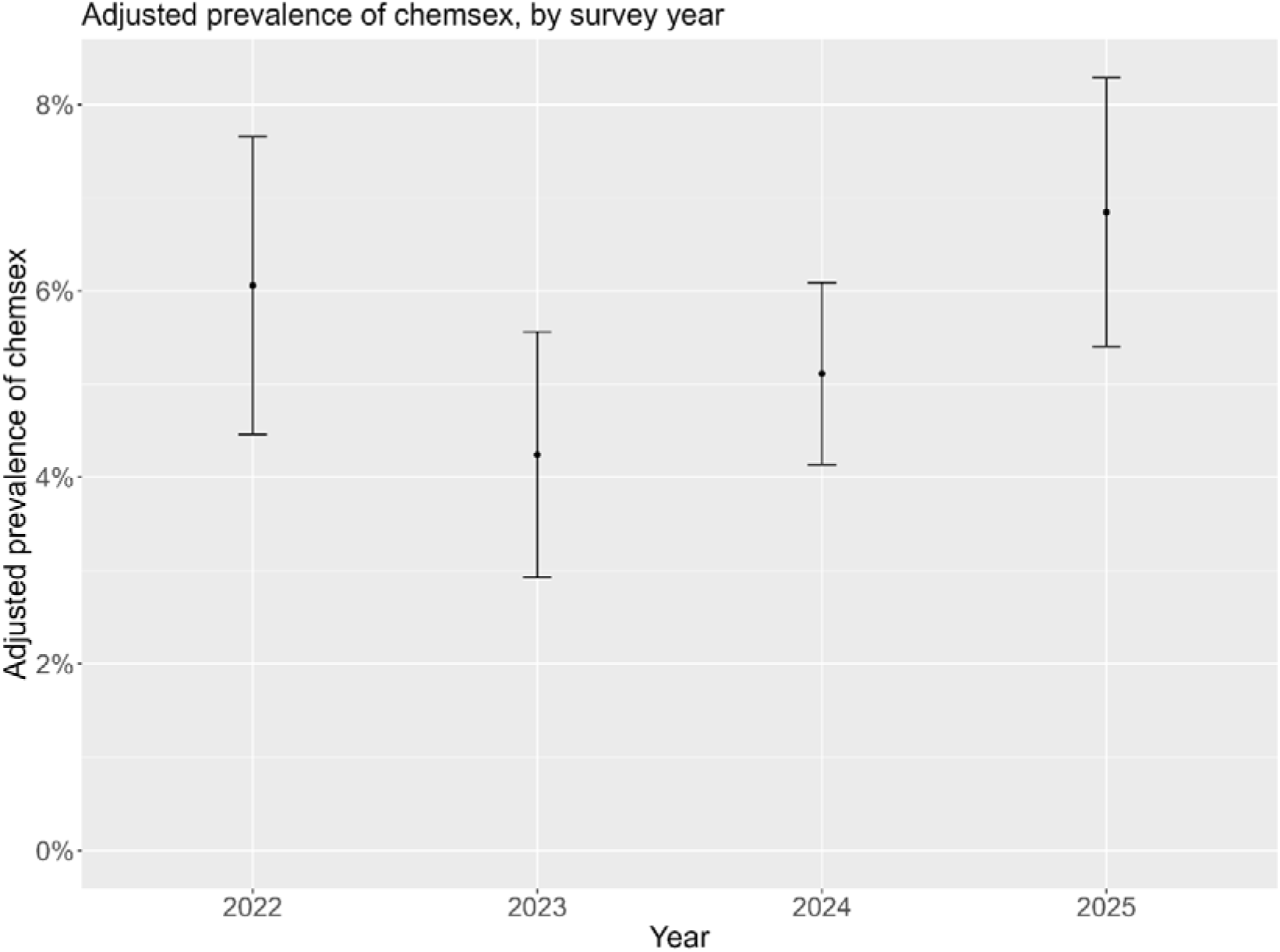
adjusted prevalence of chemsex, by survey year. Bars represent 95% confidence intervals.

### Factors associated with recent chemsex

In univariable and multivariable analyses, participants were more likely to report recent chemsex if they were gay (aOR=1.57 versus any other sexual orientation; 95%CI 1.11-2.29, p=0.01), of older age (35-59 versus 16-34 years; p<0.001), unemployed (aOR=3.08 vs employed full-time; 95%CI 2.19-4.26, p<0.001), living with HIV (aOR=3.16 versus not living with HIV/unknown HIV status; 95%CI 2.43-4.09, p<0.001) and/or had recently used HIV PrEP (aOR=3.79 vs no recent HIV PrEP use; 95%CI 2.63-5.56, p<0.001) or a sexual health service (aOR=2.23 vs no recent SHS; 95%CI 1.76-2.83, p<0.001).

## Discussion

In this large study of GBMSM in England, we found low and broadly stable recent chemsex prevalence between 4-7% in the 2022-2025 period, with small year-on-year fluctuations. Recent chemsex in England was associated with older age (35-59 vs 16-34 years), identifying as gay, living with HIV, unemployment, recent HIV PrEP use, and recent sexual health service use.

Although there was statistical evidence of an association between survey year and chemsex in the main analysis this was not robust to sensitivity analysis when adjusted for a measure of sexual risk. The adjusted point prevalence estimate for 2022 was also lower in sensitivity analysis. Combined, this suggests the fluctuations in chemsex prevalence over time are more likely due to differences in the RiiSH study participants’ characteristics over this period, rather than a true change in chemsex prevalence between 2022-2025.

Heterogeneity in the definition of chemsex – particularly of what drugs it includes and the length of the recall period – and the population surveyed across studies, limit the validity of comparisons with other chemsex prevalence estimates (Maxwell et al., 2019). However, our estimates were similar to the 6.5% prevalence of chemsex-associated drug use in the past 4 weeks amongst UK-based men who have sex with men(MSM) reported by the 2017-2018 European MSM Internet Survey (Avallone et al., 2025) and were slightly higher than the 3.3% chemsex prevalence reported in the past 3 months amongst people living with HIV attending an HIV clinic in Brighton between 2019-2020 (Adler et al., 2022). Analysis of prevalence over a longer time period will identify any emerging trends with greater certainty.

Higher odds of chemsex amongst recent users of PrEP and SHS indicate high engagement with prevention and healthcare, and supports recent findings that PrEP initiation and high PrEP adherence amongst GBMSM who engage in chemsex is driven by a desire to reduce HIV risk (Maxwell et al., 2022). Given the higher risk of acquiring sexually transmitted infections with chemsex, participants may have greater need for SHS to access treatment. Sexual health service use was higher amongst RiiSH participants reporting chemsex in 2016-2017 (Blomquist et al., 2020), and other literature also report associations between SHS use and chemsex (Hibbert et al., 2019). Routine provision of PrEP in SHS started in October 2020 so the association between SHS and chemsex may reflect consultations related to PrEP, or participants’ broader attempts to maintain sexual health as part of a chemsex risk reduction strategy (Maxwell et al., 2019). Previous studies in England have found that men engaged in chemsex would prefer specialist support to be provided in sexual health clinics (Tomkins et al., 2019), and our findings would endorse this.

Only 1.6% (91/5728) of study participants reported recent injecting drug use, but the vast majority (78/91; 86%) of these individuals also reported recent chemsex. This strong link between recent injecting drug use and recent chemsex suggests that in this small group of participants, injecting chemsex-associated drugs – or ‘slamming’ – may be one way in which they engage in chemsex (Scheibein et al., 2021), and/or that one may predispose to the other. Although it is thought that chemsex-associated drugs are mainly consumed orally or nasally, there is concern of an increasing trend in the frequency of slamming practice across men who have sex with men internationally (Schreck et al., 2021). In our study, a quarter of participants reporting recent chemsex also reported recent injecting drug use, so were potentially at additional risk of blood-borne infections if sharing injecting equipment. Understanding the evolving epidemiology of slamming is an important area for further research, particularly given the small number of participants reporting recent injecting drug use in this study, to meet the health needs of people engaging in slamming.

### Strengths

Chemsex can be a sensitive and stigmatising topic to research, which adds challenges to the estimation of its prevalence. Using an online survey that collected no personal identifiers helped to mitigate social desirability bias and limiting the recall period to the last 3-4 months reduced recall bias. Although RiiSH is a convenience sample, its strength is the large numbers of GBMSM participants it recruits annually (ranging from 800-2300 in the four years included in this study). Other national surveys of sexual health which recruit probabilistic samples, such as the National Survey of Sexual Attitudes and Lifestyles (Natsal), are more representative of the whole population, but are limited in the precision of any GBMSM-specific estimates due to the lower absolute number (n=190 in Natsal-3) of GBMSM in their studies (Mercer et al., 2016; Prah et al., 2016).

### Limitations

The RiiSH study recruits by convenience, rather than probability sampling. Although RiiSH participants are recruited from different sources, our results are not necessarily generalisable to the wider GBMSM population in England, as they exclude individuals who do not use geospatial networking or social media apps, or who do not access the internet. Additionally, it has previously been reported that compared with probability samples, GBMSM participating in convenience sample surveys tend to report more sexual risk behaviours, so the true prevalence of chemsex across all GBMSM in England may be lower than we estimate (Prah et al., 2016).

The RiiSH survey does not currently ask participants who report injecting drug use to specify the drugs they have injected, so it is not possible to measure the prevalence of slamming or to know if participants who report recent injecting drug use are injecting chemsex-associated or other drugs.

We have hypothesised associations between variables based on literature and expert discussion as outlined in the DAG, but it is possible that reverse causation explains observed associations due to the cross-sectional design of RiiSH. For example, where we have represented sexual health service use as preceding chemsex in the DAG, service use may have occurred after chemsex for the purposes of interventions such as HIV, hepatitis C and STI testing.

## Conclusion

Chemsex was reported by a minority of GBMSM, with a low and broadly stable prevalence between 4-7% in the 2022-2025 period.

HIV PrEP and SHS use was associated with chemsex suggesting that services are reaching people with greatest health needs. Our findings support the integration of specialist chemsex support within SHS.

## Acknowledgements

CRediT statement:

- Jin-Min Yuan: conceptualisation; methodology; formal analysis; investigation; writing – original draft
- Laura Viviani: conceptualisation; methodology; writing – review & editing
- Dolores Mullen: data curation; writing – review & editing
- Dana Ogaz: conceptualisation; methodology; writing – review & editing
- Hamish Mohammed: conceptualisation; methodology; writing – review & editing; supervision

We thank Dr Gavin Dabrera and Dr Helen Crabbe for reviewing the study protocol and manuscript.

## Data availability statement

The authors cannot make the underlying dataset publicly available for ethical and legal reasons.

