## APPENDICES for "Trends in, and factors associated with, chemsex in a community sample of gay, bisexual, and other men who have sex with men in England, 2022-2025"

**Appendix 1: search strategy for the rapid literature review**

A rapid literature review was undertaken of existing associations between independent variables and chemsex, to inform the potential variables and confounders that should be considered for inclusion in the multivariable model.

Using the following search strategy in PubMed with no restriction on the year of publication, 279 papers were identified, of which 96 were identified as potentially relevant following title review, 44 were kept after abstract review, and 43 included in the final literature review:

(factor* OR demographic* OR correlate* OR characteristic*) AND (chemsex*)

Only published peer-reviewed articles were included in the review.

**Appendix 2: all variables directly collected by, or derived from, the RiiSH survey across all survey rounds between 2022-2025**

- Demographics
  - Age
  - Sex assigned at birth/gender identity
  - Sexual orientation
  - Ethnicity
  - Country of birth
  - Region of residence
  - Educational level
  - Employment
  - Living situation
  - Financial situation
  - Use of social networking/dating apps
- HIV testing
  - Living with HIV (Y/N)
  - HIV testing recency
- PrEP
  - PrEP ever use
  - PrEP last used (which year)
  - PrEP use frequency since August 2024 (past 3 months)
- STI prophylaxis
  - DoxyPEP ever use
  - DoxyPEP haven’t used but considered using
  - DoxyPEP last used (which year grouping)
- STI (but not HIV) testing
  - STI (but not HIV) ever test
  - STI (but not HIV) last test (which year grouping)
  - STI (but not HIV) test frequency in last year
  - STIs (but not HIV) in the past 3 months
- Vaccination
  - Hep A vaccine (number of doses)
  - Hep B vaccine (number of doses)
  - HPV vaccine (number of doses)
  - Mpox vaccine (number of doses)
- Changed behaviours to avoid mpox infection
- Sexual health symptoms
  - Had symptoms in past 3 months
  - Seen healthcare professional about the symptoms in the past 3 months?
- Sexual health services
  - Ever visited
  - Last visit (which year grouping)
- Chemsex
  - Ever taken chemsex-associated drugs
  - When last taken chemsex-associated drugs (which year grouping)
  - Proportion of sex done after taking chemsex drugs, in past 3 months
- Injecting drugs
  - Ever injected drugs (apart from medicines or anabolic steroids)
  - Injected drugs (apart from medicines or anabolic steroids) in past 3 months
- Alcohol and sex
  - Proportion of sex whilst under influence of alcohol, in past year
  - Frequency of drinking ≥8 units of alcohol on a single occasion (binge, for men)
- Sexual satisfaction and personal wellbeing
  - Satisfaction with sex life, in past 3 months
  - Satisfaction with life
  - Life activities worthwhile?
  - Happiness yesterday
  - Anxiety yesterday
  - Physical health conditions lasting/expected to last ≥1 year
  - Physical health conditions limiting day-to-day activities
  - Mental health conditions lasting/expected to last ≥1 year
  - Mental health conditions limiting day-to-day activities
  - Health condition/disability/medication affecting sexual activity/enjoyment, in last year
  - How health condition/disability/medication has affected sexual activity/enjoyment (positive or negative way)
- Sexual relationships with men
  - Physical sexual contact with how many men, in past 3 months
  - Number of new physical sex partners (men) in past 3 months
  - Anal sex with a man, in past 3 months
  - Anal sex with how many men in past 3 months
  - Condomless anal sex in last 3 months
  - Condomless anal sex with how many men in past 3 months
  - Number of HIV concordant men they’ve had condomless anal sex with in past 3 months
  - Methods for meeting/contacting male partners for physical sexual contact in past 3 months
- Sexual relationships with women
  - Physical sexual contact with how many women, in past 3 months
  - Number of new physical sex partners (women) in past 3 months
  - Vaginal or anal sex with a woman in past 3 months
  - Condomless vaginal or anal sex with a woman in past 3 months
  - Condomless vaginal or anal sex with how many women in past 3 months
  - Number of HIV concordant women they’ve had condomless vaginal or anal sex with in past 3 months

**Appendix 3: directed acyclic graph (DAG) and list of confounder-adjustment set for each independent variable of interest**

The construction of the following directed acyclic graph was informed by a rapid literature review and expert discussion.

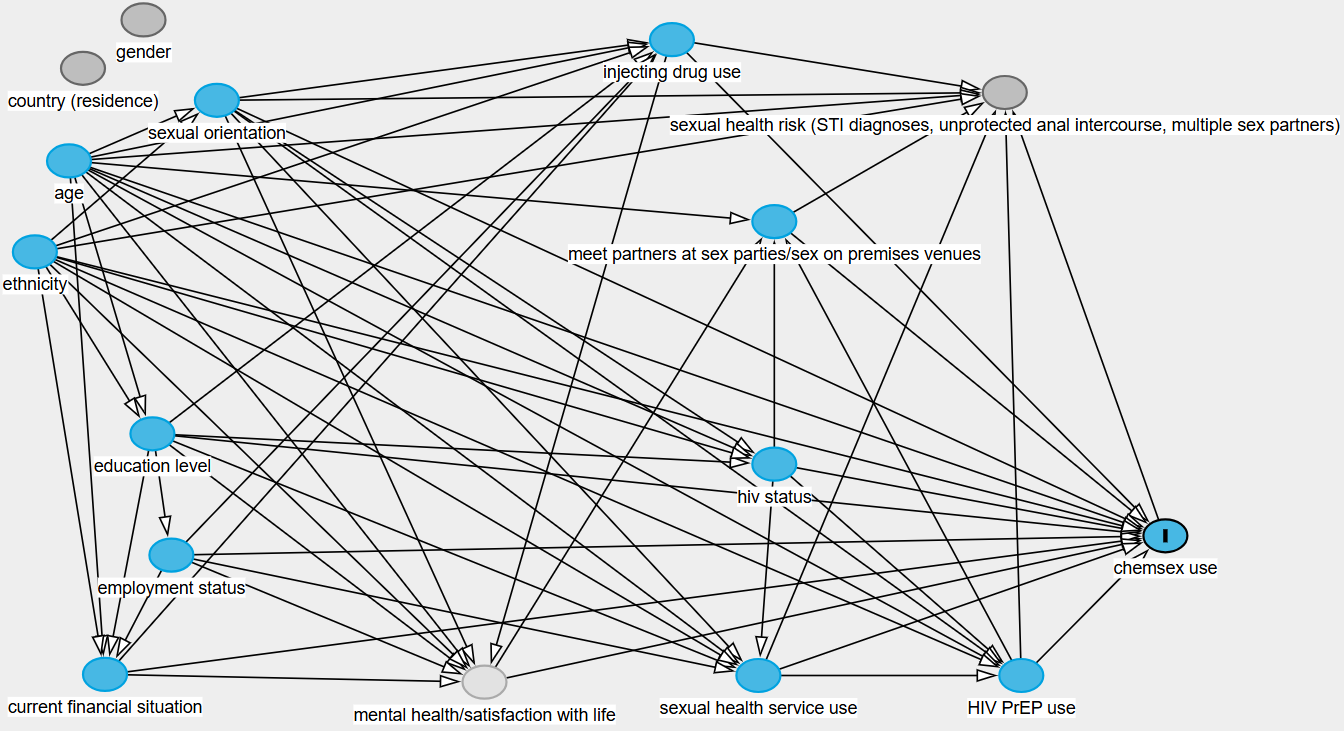

To obtain unbiased estimates of the total effect of each variable of interest on the practice of recent chemsex, we used this DAG to identify the minimum set of confounders that needed to be adjusted for in the multivariable model for each variable of interest (i.e. the minimally-sufficient adjustment set). For example, for ‘employment status’, the minimally-sufficient adjustment set from the DAG is “education level”. As age is considered a baseline covariate, the final set of covariates in the multivariable model is: “education level” and “age”. The following table describes the minimally-sufficient adjustment set for each variable of interest.

*Table: list of confounders that have been adjusted for in the multivariable model for each variable of interest, as derived from the directed acyclic graph.*

| **Variable of interest** | **Confounders adjusted for in each multivariable model (plus age as baseline covariate)** |
| --- | --- |
| Age | None |
| Education level | Age, ethnicity |
| Employment status | Age, education level |
| Sexual orientation | Age, ethnicity |
| Recent PrEP use | Age, ethnicity, HIV status, recent sexual health service use, sexual orientation |
| Recent sexual health service use | Age, ethnicity, education level, employment status, HIV status, sexual orientation |
| HIV status | Age, ethnicity, education level, sexual orientation |

**Appendix 4: sensitivity analysis – trends in chemsex prevalence**

In this model, survey year was the variable of interest and we adjusted for recruitment source, age, and the number of condomless anal intercourse partners participants reported having sex with since August of each survey year as a measure of sexual risk. After adjustment, year was not significantly associated with recent chemsex (*p* = 0.18).

Adjusted chemsex point prevalence estimates fluctuated, from 5.7% (95%CI 4.3-7.1) in 2022 to 4.9% (95%CI 3.5-6.4) in 2023, 5.1% (95%CI 4.1-6.0) in 2024 and 6.7% (95%CI 5.3-8.0) in 2025

Table: multivariable analysis of chemsex prevalence. *Adjusted for recruitment source, age and number of recent male condomless anal intercourse partners, with 2022 as the base (reference) year

| **Characteristic** | **Adjusted risk ratio*** | **95% CI** | **p-value** |
| --- | --- | --- | --- |
| Year |  |  | 0.18 |
| 2022 | — | — |  |
| 2023 | 0.87 | 0.59, 1.25 | 0.5 |
| 2024 | 0.89 | 0.64, 1.25 | 0.5 |
| 2025 | 1.17 | 0.83, 1.66 | 0.4 |

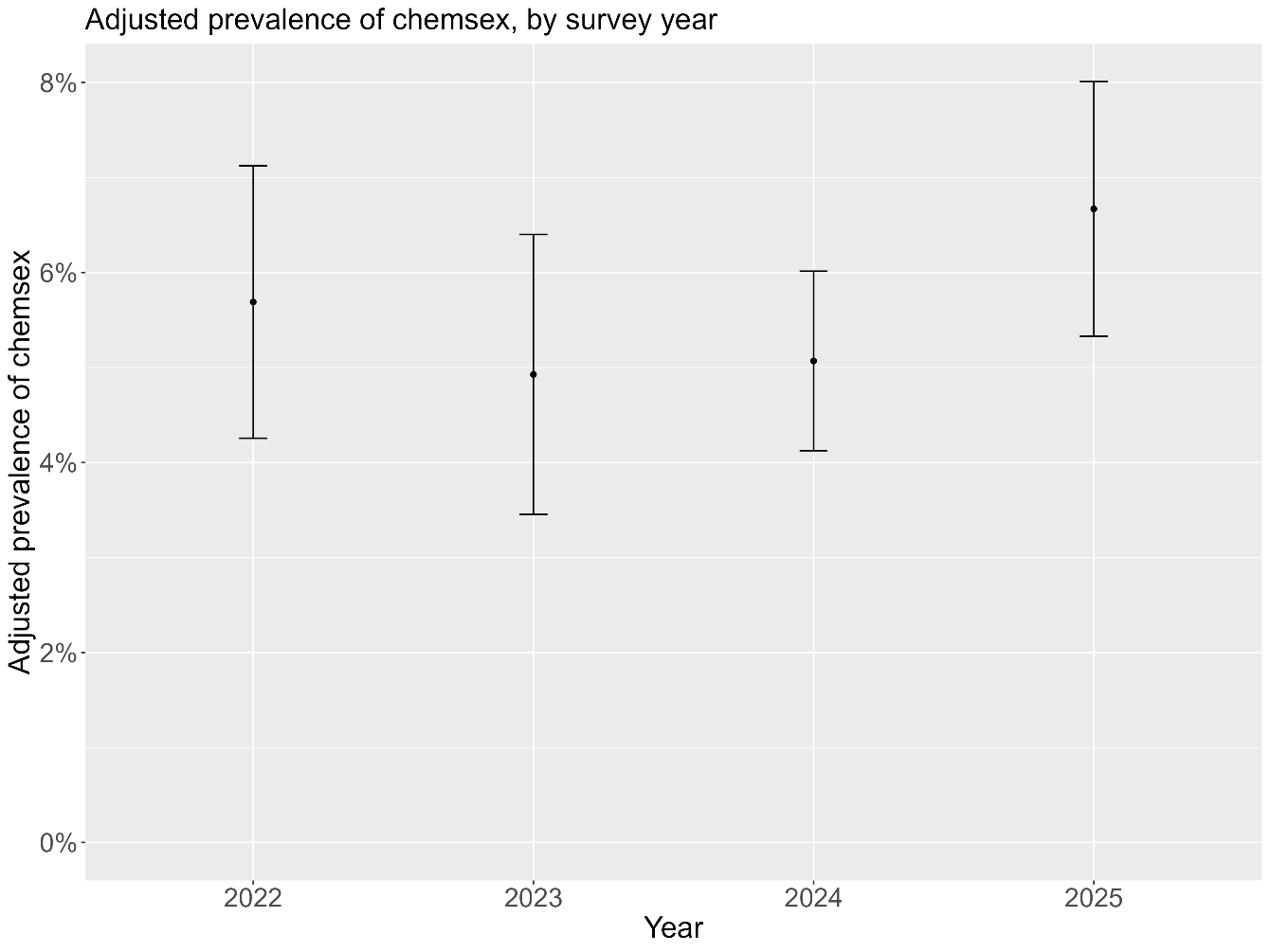

Figure: adjusted prevalence of chemsex, by survey year. Bars represent 95% confidence intervals.
